# Blood DNA methylation levels of the oxytocin promoter predict conversion from mild cognitive impairment to dementia in females within a clinical cohort of cognitive complaints

**DOI:** 10.1101/2024.08.28.24312742

**Authors:** Philippos Koulousakis, Rick Reijnders, Inez Ramakers, Frans Verhey, Tim Vanmierlo, Daniël L.A. van den Hove, Renzo J.M. Riemens

## Abstract

Recent studies have highlighted the role of oxytocin (OXT) in Alzheimer’s disease (AD) dementia and demonstrated its potential as a therapeutic target to reverse cognitive impairment and mitigate AD pathology. Epigenetic dysregulation of *OXT* has been identified in brain tissue from AD patients, and DNA methylation levels of the exact same locus in the blood of healthy aged individuals have shown predictive biomarker value for conversion to AD. Building on these insights, we investigated the DNA methylation status of the *OXT* promoter in blood in a prospective cohort of consecutive patients from the BioBank Alzheimer Center Limburg (BBACL). This cohort included males and females suffering from subjective cognitive decline (SCD), mild cognitive impairment (MCI), and dementia. Our findings revealed that DNA methylation levels of the *OXT* promoter at baseline predict the conversion from MCI to dementia in female participants. In addition to discovering differences in the *OXT* promoter related to sex, we also observed alterations associated with aging, alcohol consumption, and smoking. Overall, our findings underscore the implications of *OXT* and its DNA methylation changes in blood within the context of dementia.

## Introduction

Oxytocin (OXT) is a neuropeptide hormone that acts as both a neuromodulator and a neurotransmitter, involved in many central processes in the brain and periphery [1]. OXT is produced in two hypothalamic nuclei: the paraventricular nucleus (PVN), and the supraoptic nucleus (SON) [2]. The projections from these nuclei and the receptors for OXT are widely distributed throughout the brain, and innervate regions such as the forebrain, hippocampus, amygdala, the perirhinal- and entorhinal cortices, and the supramammillary nucleus [3–5], among others. From the posterior pituitary, OXT is released into the bloodstream where it exerts its peripheral effects [5].

Studies have identified roles for OXT in social bonding, intimacy, anxiety, and cognitive processes, including memory [6–8]. OXT acts within the central nervous system to regulate these behavioral processes, contrasting its well-known peripheral role in childbirth and lactation [1]. Furthermore, roles for OXT in mediating the stress response [4], inflammation [9], and nociception [10] have been described as well. For further details on the physiological roles of OXT, its receptors and its circuitry, we would like to refer the reader to two related reviews [5, 11].

Beyond its physiological roles, research has established a connection between OXT and various human illnesses, including dementia [5, 7]. A potential role for OXT has become evident based on *e.g.*, the identified link between social isolation, amyloid beta (Aβ) burden, and the risk of developing Alzheimer’s disease (AD) dementia [5, 12]. Furthermore, altered OXT concentrations and signaling have been observed in brain areas involved in cognitive functioning in AD dementia patients [13, 14]. In addition, studies in rodents have reported that OXT administration can reverse Aβ-induced cognitive impairments [15–17] and that it has potential therapeutic effects by suppressing acetylcholinesterase activity, reducing hippocampal Aβ deposition, and lowering tau levels [7].

Additionally, altered gene expression profiles in OXT signaling have been observed when comparing bloods samples from patients with mild cognitive impairment (MCI) and AD dementia [18]. Previous research has also identified epigenetic differences in *OXT* in AD dementia patients in the middle temporal gyrus (MTG) [19] and in the superior temporal gyrus (STG) [20]. Strikingly, DNA methylation changes of the *OXT* promoter in the same locus as identified in the brain have been associated with conversion to AD dementia in blood of healthy aged individuals, giving blood DNA methylation levels of *OXT* predictive biomarker value [19]. Taken together, these studies suggest that OXT might be a target for future therapeutic strategies and the development of biomarker assays in AD dementia.

The potential of using *OXT* DNA methylation in clinical blood samples as a biomarker for predicting conversion to AD dementia is particularly compelling, owing to the method’s ease and minimal invasiveness. Furthermore, DNA methylation changes in *OXT* have been documented not only in blood samples but also in different brain regions, emphasizing its significant role in the disease [21]. First however, it is crucial to further evaluate both the robustness of these findings and the causality of the observed epigenetic associations within *OXT*. This is essential to confirm the relevance of these alterations in AD pathology and to determine the specificity of the epigenetic association within the context of dementia. Thus, replication and validation of these findings in different dementia patient cohorts are necessary.

In the present study, we aimed to investigate whether differential DNA methylation levels in the *OXT* promoter could be detected in blood samples from a clinical cohort with cognitive complaints, sourced from the BioBank Alzheimer Center Limburg (BBACL). This cohort included individuals with subjective cognitive decline (SCD), MCI, and dementia (Figure 1). Specifically, we sought to determine whether there were distinct DNA methylation patterns in the *OXT* promoter among these three groups. In view of the longitudinal design of our cohort study, we also aimed to assess whether the DNA methylation status of the *OXT* promoter at baseline could predict conversion to dementia in MCI patients.

**Figure 1.**
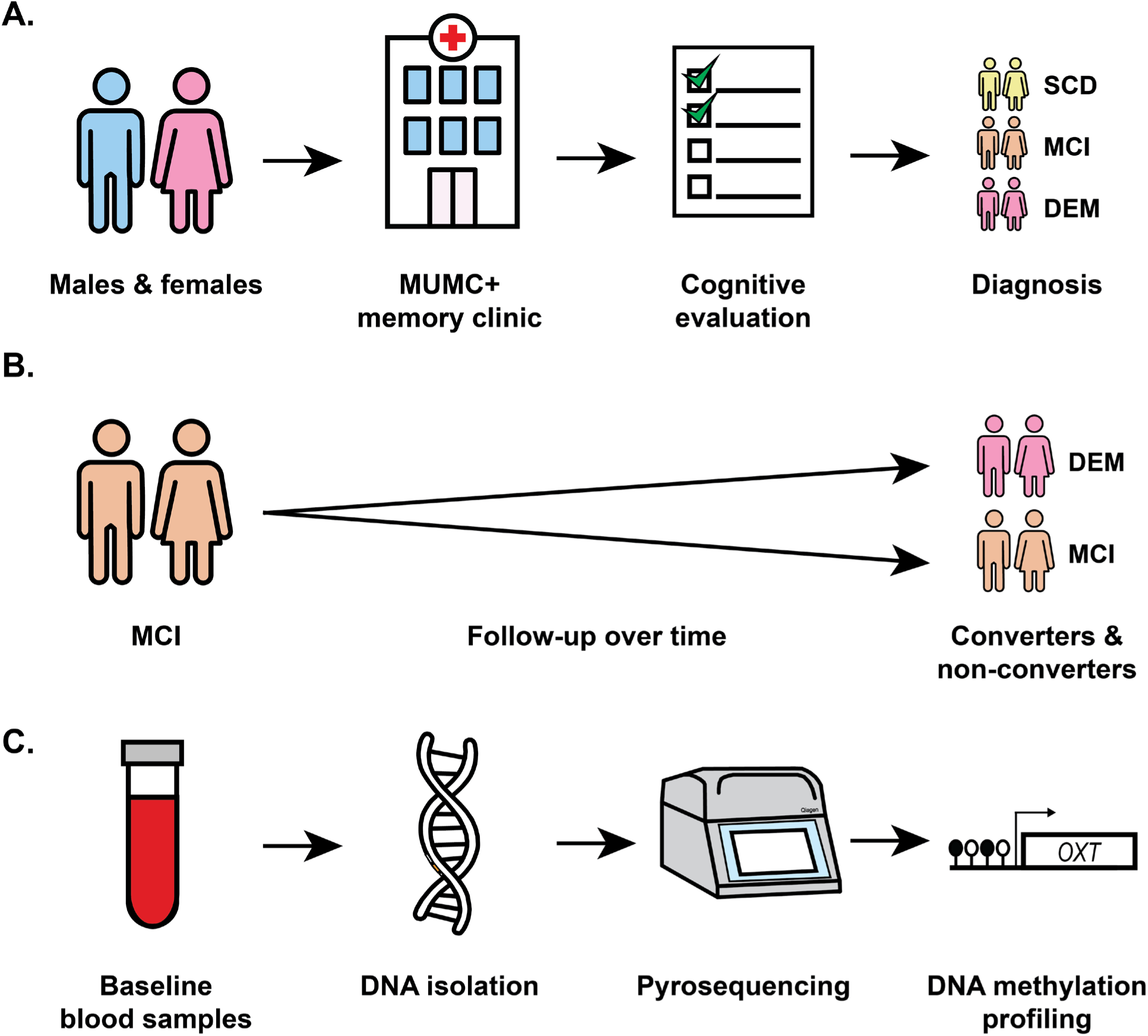
Visual representation of the workflow from patient recruitment and sample collection to processing. **A.** Patients referred to the memory clinic at Maastricht University Medical Center + (MUMC+) underwent cognitive evaluations and were diagnosed with subjective cognitive decline (SCD), mild cognitive impairment (MCI), or dementia (DEM). **B.** MCI patients were followed up over time, and some subsequently developed dementia (converters), while others did not develop dementia (non-converters). **C.** Whole blood samples at baseline were collected, the buffy coat was processed for DNA isolation, the DNA was bisulfite-converted, and the *OXT* promoter was pyrosequenced.

## Materials and methods

### Study sample description

For this study, 260 individuals (147 males, 113 females) from the BBACL cohort were selected, including 39 individuals with SCD (33 males, 6 females), 168 patients with MCI (89 males, 79 females), and 53 patients with dementia (25 males, 28 females). The BBACL cohort comprises individuals who were referred to the Memory Clinic for cognitive evaluations at Maastricht University Medical Center + (MUMC+) in Maastricht, the Netherlands. Eligibility criteria required participants to have a clinical dementia rating scale (CDR) score between 0 and 1 and a Mini-Mental State Examination (MMSE) score of ≥ 20. Exclusion criteria at baseline included non-degenerative neurological conditions such as Normal Pressure Hydrocephalus, Huntington’s disease, brain cancer, epilepsy, encephalitis, transient ischemic attack (TIA), or cerebrovascular accident (CVA) within the past two years. Additionally, individuals with a recent TIA or CVA accompanied by cognitive decline, patients with a history of psychiatric disorders, current major depressive disorder within the last 12 months (DSM IV), or alcohol abuse, were also excluded. All participants underwent a physical, cognitive, and neuropsychiatric evaluation, and whole blood samples were collected. Among the 168 MCI patients, 88 developed dementia (44 males, 44 females), while 80 did not develop dementia (45 males, 35 females) 76 months after baseline. The BBACL study protocol was approved by the local ethics committee (METC 15-4-100) at MUMC+ and all participants provided written informed consent.

### DNA isolation

The QIAsymphony DSP DNA Midi kit (Qiagen, Hilden, Germany) was used to isolate DNA from whole blood buffy coat according to the manufacturer’s instructions. DNA concentrations were measured using the Qubit dsDNA BR Assay Kit (Thermo Fisher Scientific, Waltham, MA, USA) and the Qubit 2.0 Fluorometer (Thermo Fisher Scientific, Waltham, MA, USA), following the manufacturer’s protocol. The isolated DNA was stored at -80°C until further processing.

### Bisulfite treatment

Bisulfite treatment was performed on 200 ng of DNA per blood sample using the EZ-96 DNA Methylation-Gold kit (D5008, Zymo Research, Irvine, CA, USA) according to the manufacturer’s instructions, with the exception of eluting the bisulfite-converted samples in 20µL of elution buffer. This resulted in a final concentration of 10ng/µL bisulfite-converted DNA. Samples were randomized over the different plates to avoid batch effects.

### Polymerase chain reaction

Polymerase chain reaction (PCR) and pyrosequencing primers for the *OXT* promoter were designed using the PyroMark Assay Design software version 2.0.1.15, based on the Ensembl Genome Browser GRCh37 assemble database (Supplementary Table 1). PCR reactions were performed using the FastStart Taq DNA Polymerase, dNTPack (Roche, Basel, Switzerland). Thermocycler settings included an initial denaturation step at 95°C for 5 minutes, followed by 45 cycles at 95°C for 30 seconds, 62°C for 30 seconds, and 72°C for 30 seconds, with a final elongation step of 72°C for 1 minute. Reactions comprised 2.5 μl PCR buffer (10X) with 20 mM MgCl2, 0.5 μl 10 mM dNTP mix, 1 μl of each primer (5 μM stock), 1μl of bisulfite-converted DNA, and 0.2 μl (5 U/μl) FastStart Taq DNA Polymerase (Roche Diagnostics GmbH, Mannheim, Germany) in a total volume of 25 μl. PCR products were size-fractionated on a 2% agarose gel in Tris-Acetate-EDTA (TAE) buffer.

### Pyrosequencing

The DNA methylation (5-methylcytosine, 5-mC) levels of the *OXT* promoter were quantified for 8 cytosine-phosphate-guanine (CpG) sites. The targeted region spanned from 3052247 to 3052237 on chromosome 20 (Ensembl GRCh37 assembly). Pyrosequencing was performed using the PyroMark Q48 Autoprep system with the Pyro Q48 Autroprep 2.4.2 software (Qiagen, Hilden, Germany), and the PyroMark Q48 Advanced Reagents following the manufacturer’s instructions. Samples were randomized over different plates to avoid batch effects. Assay sensitivity was confirmed using DNA standards from the EpiTect PCR Control DNA Set (Qiagen). Pyrosequencing data of CpG sites that did not meet the quality control (QC) assessment by the Pyro Q-CpG 1.0.9 software were excluded pairwise from downstream analyses.

### Experimental design

DNA methylation levels in the *OXT* promoter were initially analyzed using linear regression modeling to compare the three experimental groups (SCD, MCI, and dementia), while adjusting for covariates known to impact methylation, including age [22], sex [23], smoking [24], and alcohol consumption [25]. Subsequent contrast analyses were conducted to separately compare individuals with SCD to those with MCI, SCD to dementia patients, and MCI to dementia patients. Additionally, we examined differential methylation levels in the *OXT* promoter at baseline between dementia converters and non-converters within the MCI group. In cases where significant sex effects were identified, further analyses were stratified by sex, separating male and female participants. Furthermore, for each analysis where two or more variables were significant for the same CpG site, interaction effects were tested to evaluate their combined influence.

### Statistical analysis

All computational and statistical analyses were conducted using R (version 3.3.2) and Rstudio (version 1.0.136). Linear regression modeling for differential DNA methylation levels of the *OXT* promoter was performed based on diagnosis (SCD, MCI, and dementia) and conversion to dementia (converters and non-converters within the MCI group). To reduce data variance, age, sex, alcohol consumption, and smoking were included as covariates in the linear regression models. Alcohol consumption and smoking were characterized as dichotomous variables, either indicating a history or current use (alcohol; smoker), or indicating no history and no current use (no alcohol; no smoker). A *p*-value < 0.05 was considered nominally significant, whereas a false-discovery rate (FDR)-adjusted *p-*value < 0.05 was considered statistically significant. FDR correction was applied using the Benjamini-Hochberg procedure. Cases with missing demographic data were excluded list-wise. After QC assessment using the Pyro Q-CpG 1.0.9 software and subsequent data curation, the final dataset included 30 individuals with SCD, 125 patients with MCI, and 37 dementia patients (Table 1). Among the 125 patients with MCI, 63 developed dementia, while 62 did not develop dementia.

**Table 1.** Demographics of studied cohort.

|  | **SCD** | **MCI** | **Dementia** |
| --- | --- | --- | --- |
| **Sample size (%)** |  |  |  |
| Male + Female | 30 (15.63) | 125 (65.10) | 37 (19.27) |
| Male | 25 (83.33) | 70 (56.00) | 18 (48.65) |
| Female | 5 (16.67) | 55 (44.00) | 19 (51.35) |
| **Age (years) ± SD** |  |  |  |
| Male + Female | 59.37 ± 10.03 | 72.13 ± 7.44 | 72.97 ± 8.25 |
| Male | 59.52 ± 10.84 | 72.49 ± 7.29 | 71.11 ± 6.76 |
| Female | 58.6 ± 4.93 | 71.67 ± 7.66 | 74.74 ± 9.28 |
| **Dementia converter (%) / non-converter (%)** |  |  |  |
| Male + Female | N.A. | 63 (50.40) / 62 (49.60) | N.A. |
| Male |  | 36 (51.43) / 34 (48.57) |  |
| Female |  | 27 (49.09) / 28 (50.91) |  |
| **Alcohol (%) / no alcohol (%)** |  |  |  |
| Male + Female | 25 (83.33) / 5 (16.67) | 95 (73.00) / 30 (24.00) | 30 (81.08) / 7 (18.91) |
| Male | 21 (84.00) / 4 (16.00) | 59 (84.29) / 11 (15.17) | 16 (88.89) / 2 (11.11) |
| Female | 4 (80.00) / 1 (20.00) | 36 (65.45) / 19 (34.55) | 14 (73.98) / 5 (26.32) |
| **Smoking (%) / no smoking (%)** |  |  |  |
| Male + Female | 17 (56.67) / 13 (43.33) | 69 (55.20) / 56 (44.80) | 14 (37.84) / 23 (62.16) |
| Male | 13 (52.00) / 12 (48.00) | 44 (62.86) / 26 (37.14) | 9 (50.00) / 9 (50.00) |
| Female | 4 (80.00) / 1 (20.00) | 25 (45.45) / 30 (54.55) | 5 (26.32) / 14 (73.68) |

SCD = subjective cognitive decline; MCI = mild cognitive impairment; SD = standard deviation. The sample size for each experimental group is presented as the number, with the percentage of the total cohort shown in parentheses. The number and percentage of males and females are provided for each individual experimental group.

## Results

### Comparison of DNA methylation levels in the OXT promoter across SCD, MCI, and dementia groups

Linear regression analysis exploring the association between DNA methylation levels in the *OXT* promoter and diagnostic groups (SCD, MCI, and dementia) identified nominally significant age-related effects at CpG sites 1 (*p* = 0.029), 2 (*p* = 0.007), 3 (*p* = 0.009), 4 (*p* = 0.036) and 5 (*p* = 0.033; Supplementary Figure 1A-E). Following multiple testing correction, CpG sites 2 (FDR-adjusted *p* = 0.035) and 3 (FDR-adjusted *p* = 0.035) remained significant, indicating that DNA hypomethylation is observed with increasing age. Additionally, nominally significant effects associated with sex were observed for CpG sites 1 (*p* = 0.003), 2 (*p* = 0.006), 3 (*p* = 0.033), 4 (*p* = 0.026), 5 (*p* = 0.046), 7 (*p* = 0.004), and 8 (*p* = 0.010; Supplementary Figure 5A). Following FDR correction, CpG sites 1 (FDR-adjusted *p* = 0.015), 2 (FDR-adjusted *p* = 0.016), 3 (FDR-adjusted *p* = 0.044), 4 (FDR-adjusted *p* = 0.042), 7 (FDR-adjusted *p* = 0.015), and 8 (FDR-adjusted *p* = 0.021) remained statistically significant. DNA hypermethylation at these sites was observed in females compared to males. An interaction test examining the combined effects of age and sex on DNA methylation levels of CpG sites 2 and 3 did not reveal a statistically significant interaction effect (data not shown). Furthermore, no significant associations were found for the primary predictor, diagnosis (Figure 2A), or for the covariates alcohol consumption (Supplementary Figure 6A) and smoking (Supplementary Figure 7A) across the SCD, MCI, and dementia groups.

**Figure 2.**
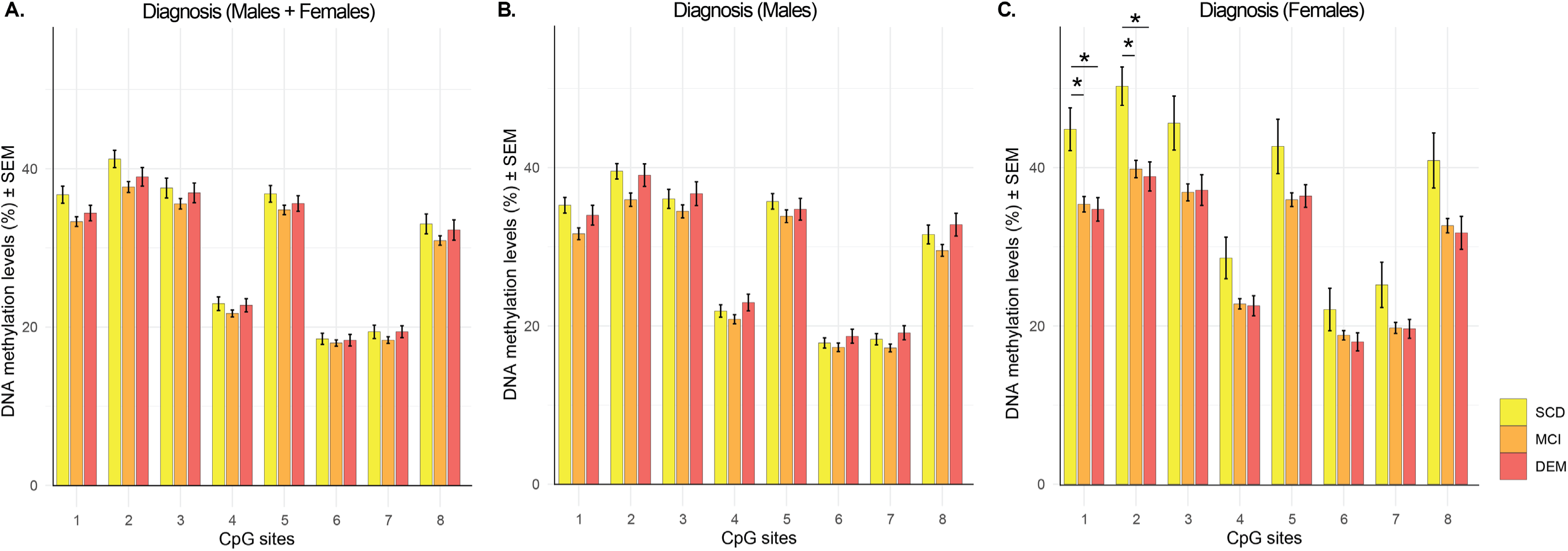
Average CpG site DNA methylation levels by diagnosis. The bar charts show the average cytosine-phosphate-guanine (CpG) site DNA methylation levels in the *OXT* promoter ± the standard error of the mean (SEM). These averages are presented for the diagnostic groups subjective cognitive decline (SCD), mild cognitive impairment (MCI), or dementia (DEM), both for males and females combined, as well as separately. Nominal significance (*p* < 0.05) related to the primary predictor diagnosis is indicated with an asterisk (*), whereas FDR-adjusted statistical significance (FDR-adjusted *p* < 0.05), if applicable, is indicated with an asterisk followed by “FDR” (*FDR).

Stratified analyses by sex revealed nominally significant associations between DNA methylation levels in the *OXT* promoter and diagnosis in females (Figure 2C). Specifically, for CpG site 1, nominally significant differences were observed between SCD and MCI (*p* = 0.027) and between SCD and dementia (*p* = 0.025). Similarly, for CpG site 2, nominally significant differences were found between SCD and MCI (*p* = 0.034) and between SCD and dementia (*p* = 0.043). Additionally, nominally significant effects associated with age were identified for CpG sites 2 (*p* = 0.017), 3 (*p* = 0.009), 5 (*p* = 0.044), and 8 (*p* = 0.044; Supplementary Figure 1B, C, E, and H). Furthermore, nominally significant smoking-related effects were identified for CpG sites 1 (*p* = 0.009), 2 (*p* = 0.002), and 7 (*p* = 0.031; Supplementary Figure 7C). After multiple testing correction, only the associations between smoking and DNA methylation levels for CpG sites 1 (FDR-adjusted *p* = 0.037) and 2 (FDR-adjusted *p* = 0.014) remained significant. These results indicate that DNA hypomethylation at CpG sites 1 and 2 is observed in female smokers compared to female non-smokers. Across the SCD, MCI, and dementia groups, no significant associations of alcohol were identified in females (Supplementary Figure 6C).

In males, neither nominally- nor FDR-adjusted significant associations were observed for diagnosis (Figure 2B) or any of the covariates analyzed (Supplementary Figure 1, 6B, 7B).

### Comparison of DNA methylation levels in the *OXT* promoter between SCD and MCI groups

Linear regression analysis comparing DNA methylation levels in the *OXT* promoter between individuals with SCD and those with MCI revealed nominally significant age-related effects for CpG sites 2 (*p* = 0.029; Supplementary Figure 2B) and 3 (*p* = 0.048; Supplementary Figure 2C). However, these effects did not survive multiple testing correction. Additionally, nominally significant sex-associated differences were identified for CpG sites 1 (*p* = 0.003), 2 (*p* = 0.004), 3 (*p* = 0.042), 4 (*p* = 0.011), 6 (*p* = 0.019), 7 (*p* = 0.002), and 8 (*p* = 0.003; Supplementary Figure 5B). The associations between sex and DNA methylation levels in the *OXT* promoter remained significant after correcting for multiple comparisons (FDR-adjusted *p*-values: CpG site 1 = 0.008, 2 = 0.008, 3 = 0.048, 4 = 0.018, 6 = 0.026, 7 = 0.008, and 8 = 0.008). These data show that DNA hypermethylation in the *OXT* promoter at these sites is observed in females when compared to males within the SCD and MCI groups. Within these two groups, no significant effects associated with diagnosis (Figure 2A), or alcohol consumption (Supplementary Figure 6D), and smoking (Supplementary Figure 7D) were identified.

Separating the males and females for stratified analyses, identified nominally significant effects on DNA methylation levels in the *OXT* promoter related to diagnosis in females for CpG sites 1 (*p* = 0.035) and 2 (*p* = 0.045; Figure 2C). Additionally, a nominally significant age effect was found for CpG site 3 (*p* = 0.025; Supplementary Figure 2C). Furthermore, nominally significant associations between smoking and DNA methylation levels in the *OXT* promoter were observed for CpG sites 1 (*p* = 0.008) and 2 (*p* = 0.005; Supplementary Figure 7F). Only these effects associated with smoking remained significant after multiple testing correction with FDR-adjusted *p*-values of 0.032 for both CpG sites. This showed that DNA hypomethylation in the *OXT* promoter at CpG sites 1 and 2 was observed in female smokers compared to female non-smokers within the SCD and MCI groups.

In males, nominally significant effects were identified for alcohol consumption for CpG sites 4 (*p* = 0.013) and 5 (*p* = 0.024; Supplementary Figure 6E), and for smoking for CpG site 4 (*p* = 0.033; Supplementary Figure 7E). However, these associations did not remain significant after multiple testing correction. No significant effects related to diagnosis were identified among males within the SCD and MCI groups (Figure 2B). Additionally, no effect of age on DNA methylation levels was observed in males (Supplementary Figure 2).

### Comparison of DNA methylation levels in the *OXT* promoter between SCD and dementia groups

Comparing DNA methylation levels in the *OXT* promoter between individuals with SCD and patients with dementia using linear regression modeling revealed a nominally significant age effect for CpG site 5 (*p* = 0.028; Supplementary Figure 3E). Additionally, nominally significant sex effects were identified for CpG sites 1 (*p* = 0.034), 2 (*p* = 0.030) and 3 (*p* = 0.043; Supplementary Table 5C). However, none of these associations remained significant after FDR-adjustment. Furthermore, no significant associations related to diagnosis (Figure 2A), alcohol consumption (Supplementary Figure 6G) or smoking (Supplementary Figure 7G) were obtained.

Stratified analyses separating males and females revealed nominally significant associations in females between diagnosis and DNA methylation levels of the *OXT* promoter for CpG sites 1 (*p* = 0.047) and 2 (*p* = 0.049; Figure 2C), but these did not survive multiple testing correction. No further associations for the females with age (Supplementary Figure 3), alcohol consumption (Supplementary Figure 6I), or smoking (Supplementary Figure 7I) were identified.

In males, no nominally significant or FDR-adjusted significant effects were observed for the primary predictor, diagnosis (Figure 2B), or any of the covariates analyzed (Supplementary Figure 3, 6H, 7H) when comparing the SCD and dementia groups.

### Comparison of DNA methylation levels in the *OXT* promoter between MCI and dementia groups

Linear regression modeling comparing DNA methylation levels in the *OXT* promoter between patients with MCI and those with dementia identified nominally significant age-related effects for CpG sites 1 (*p* = 0.019), 2 (*p* = 0.009), 3 (*p* = 0.003), 4 (*p* = 0.025), and 8 (*p* = 0.048; Supplementary Figure 4A-D, H).

Furthermore, nominally significant sex-related effects were identified for CpG sites 1 (*p* = 0.046) and 7 (*p* = 0.042; Supplementary Figure 5D), whereas a smoking-related effect was identified for CpG site 2 (*p* = 0.027; Supplementary Figure 7J). From these findings, the association related to age for CpG sites 2 (FDR-adjusted *p* = 0.035) and 3 (FDR-adjusted *p* = 0.023) remained statistically significant after FDR-adjustment. These data showed that DNA hypomethylation at CpG sites 2 and 3 is observed with increasing age for the patients within the MCI and dementia groups. No significant effects were obtained for diagnosis (Figure 2A) or alcohol consumption (Supplementary Figure 6J).

Stratified analyses by sex revealed nominally significant effects in females related to aging for CpG sites 1 (*p* = 0.038), 2 (*p* = 0.014), 3 (*p* = 0.006), 5 (*p* = 0.028), 7 (*p* = 0.037), and 8 (*p* = 0.030; Supplementary Figure 4A-C, E, G, H). After adjusting for multiple comparisons, the age-related effect for CpG site 3 (FDR-adjusted *p* = 0.045; Supplementary Figure 4C) remained significant, showing DNA hypomethylation at this site with increasing age in females within the MCI and dementia groups.

Additionally, nominally significant smoking-related effects were observed for CpG sites 1 (*p* = 0.014), 2 (*p* = 0.003), and 7 (*p* = 0.043; Supplementary Figure 7L). After FDR adjustment, the effect of smoking on CpG site 2 (FDR-adjusted *p* = 0.023) remained statistically significant, showing DNA hypomethylation at this site in female smokers compared to female non-smokers within the MCI and dementia groups. No association with diagnosis was identified for the females among these groups (Figure 2C).

In males, only a nominally significant effect related to alcohol consumption was identified for CpG site 4 (*p* = 0.030; Supplementary Figure 6K). No significant effects were identified for the main predictor, diagnosis (Figure 2B), or the other covariates smoking (Supplementary Figure 7K) and age (Supplementary Figure 4) when comparing the MCI and dementia groups.

### Comparison of DNA methylation levels in the *OXT* promoter between dementia converters and non-converters

Linear regression analysis examining the relationship between DNA methylation levels in the *OXT* promoter and conversion status from MCI to dementia revealed nominally significant associations related to sex for CpG sites 1 (*p* = 0.046), 7 (*p* = 0.027), and 8 (*p* = 0.036). However, these effects did not remain significant after multiple testing correction (data not shown). No significant findings were obtained for the effect of conversion to dementia (Figure 3A), alcohol consumption (Supplementary Figure 8A), or smoking (Supplementary Figure 9A).

**Figure 3.**
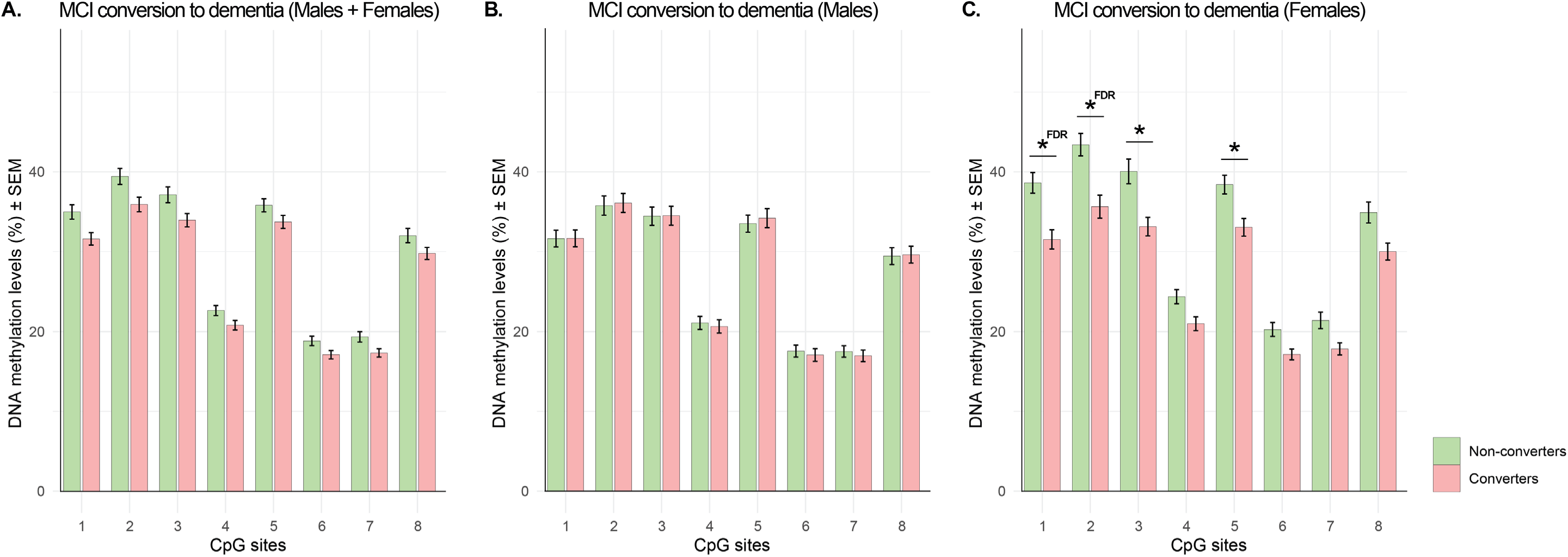
Average CpG site DNA methylation levels for dementia converters and non-converters. The bar charts show the average cytosine-phosphate-guanine (CpG) site DNA methylation levels in the *OXT* promoter ± the standard error of the mean (SEM). These averages are presented for dementia converters and non-converters in the mild cognitive impairment (MCI) group, both for males and females combined, as well as separately. Nominal significance (*p* < 0.05) related to the primary predictor conversion is indicated with an asterisk (*), whereas FDR-adjusted statistical significance (FDR-adjusted *p* < 0.05), if applicable, is indicated with an asterisk followed by “FDR” (*FDR).

Stratified analyses separating males and females revealed nominally significant effects related to conversion to dementia in females for CpG sites 1 (*p* = 0.003), 2 (*p* = 0.007), 3 (*p* = 0.045), and 5 (*p* = 0.037; Figure 3C). After applying multiple testing correction, the associations for CpG sites 1 (FDR-adjusted *p* = 0.026) and 2 (FDR-adjusted *p* = 0.027) remained statistically significant. These findings indicated that DNA hypomethylation in the *OXT* promoter at baseline is observed in females who convert to dementia compared to those who do not. Additionally, smoking-related effects were found for CpG sites 1 (*p* = 0.003) and 2 (*p* = 0.002; Supplementary Figure 9C) that remained statistically significant after multiple testing correction (FDR-adjusted *p*-values CpG sites 1 = 0.013 and 2 = 0.013). DNA hypomethylation in the female smokers was observed when compared to the female non-smokers within the MCI group for both CpG sites. An interaction test examining the combined effects of conversion and smoking on CpG sites 1 and 2 did not reveal a statistically significant interaction effect (data not shown).

In males, nominally significant effects of alcohol consumption on DNA methylation levels in the *OXT* promoter were identified for CpG sites 4 (*p* = 0.004) and 6 (*p* = 0.049), of which CpG site 4 (FDR-adjusted *p* = 0.033) remained statistically significant after FDR-adjustment. DNA hypomethylation for this CpG site was observed when comparing male alcohol consumers in the MCI group to male non-consumers. No effect of conversion (Figure 3B), age (data not shown) or smoking (Supplementary Figure 9B) were found in the males with MCI.

## Discussion

In this study, we aimed to investigate differential DNA methylation levels in the *OXT* promoter using blood samples from a clinical cohort with cognitive complaints, sourced from the BioBank Alzheimer Center Limburg (BBACL; Figure 1). This cohort included individuals with SCD, MCI, and dementia, with the advantage of longitudinal data tracking the progression of MCI patients to dementia. Our primary objectives were to delineate distinct DNA methylation patterns in the *OXT* promoter across these diagnostic groups and to determine if these methylation levels at baseline could independently predict progression to dementia in MCI patients.

Our analysis revealed a significant association between DNA methylation levels in the *OXT* promoter and dementia conversion in patients with MCI, particularly among females (Figure 3). Stratified analyses showed that hypomethylation at CpG sites 1 and 2 was observed in female dementia-converters, with these associations remaining significant after FDR correction. This demonstrates that DNA methylation levels in the *OXT* promoter could serve as potential biomarkers for predicting the conversion from MCI to dementia in female patients. The persistence of these associations after multiple testing correction reinforces the robustness of our findings, indicating a strong link between DNA methylation patterns in the *OXT* promoter and dementia conversion in females.

Consistent with our findings, altered DNA methylation signatures related to *OXT* signaling in blood have previously been associated with dementia [26]. Furthermore, a similar region of the *OXT* promoter as in our study exhibited differential methylation levels in another patient cohort predicting conversion to AD [19]. Specifically, AD converters showed a decrease in DNA methylation in blood at one of the ten CpG sites identified, while an increase was found at the other nine sites [19]. Identical to this study, DNA methylation levels were analyzed using linear regression with conversion to AD as the predictor. However, we did not compare the methylation of converters to AD dementia and healthy non-converters at a preclinical stage. In fact, our study brings a unique focus by specifically examining DNA methylation levels in patients diagnosed with MCI, representing a later stage in the disease progression, and relating this to longitudinal conversion data. The differences between the studies regarding the directionality of the effect highlight a potential role for a more dynamic regulation of DNA methylation levels in the *OXT* promoter in blood throughout dementia development and progression, meaning that methylation levels could be dependent on the stage of pathology. Such a dynamic regulation has previously also been observed in the MTG, where *OXT* hypermethylation is apparent in Braak stages 3-4, followed by hypomethylation in later stages [19].

In addition to the association with dementia conversion, we obtained nominally significant differences in DNA methylation levels across the three diagnostic categories (SCD, MCI, and dementia) for the same two CpG sites in females (Figure 2). A decrease in DNA methylation levels between SCD, and MCI and dementia at CpG sites 1 and 2 was apparent among the diagnostic groups within the females. Although these associations with diagnosis did not survive correction for multiple testing, the overlapping results for these sites across both analyses underscore their potential relevance in dementia. The consistent directionality of these associations furthermore suggests that they may be important for monitoring disease progression. However, the lack of statistically significant results after correction across the diagnostic groups indicates that these differences may not be specific enough to clearly differentiate between the different disease stages. Despite this, the observed variation in methylation patterns across different stages of dementia progression supports the potential value of these DNA modifications in the *OXT* promoter as biomarkers for predicting dementia conversion in female patients.

A primary limitation of our study that could have impacted our findings is that the BBACL cohort was not originally designed to assess differential DNA methylation levels across diagnoses. As a result, the average age varies substantially between the SCD individuals and the other diagnostic groups. Additionally, the female cohort’s relatively small sample size may have limited the statistical power to detect differences related to diagnosis. Furthermore, the BBACL cohort lacks a robust control group, given that SCD individuals from a memory clinic cohort are at an increased risk of developing MCI and subsequent dementia. To address these challenges, future studies should aim to increase the sample size to enhance statistical power and reliability. Given the broad nature of dementia diagnoses, focusing on more homogeneous subgroups within the population may reveal clearer patterns. Additionally, integrating multiple types of biomarkers—such as combining DNA modifications with other types of biomarkers like RNA sequencing data, protein expression levels or imaging data—could provide a more comprehensive understanding of their predictive power for disease progression.

Next to associations with conversion and diagnosis, our analyses revealed FDR-significant effects of smoking on DNA methylation levels at the same two CpG sites in the *OXT* promoter mentioned before. Effects of smoking on DNA methylation levels are not rare and changes have previously been identified in blood [27]. In our study, we identified effects among females in both CpG sites 1 and 2 when comparing the three diagnostic groups and when specifically examining those who progressed from MCI to dementia. Overall, DNA hypomethylation at CpG sites 1 and 2 was observed in female smokers compared to female non-smokers. However, our interaction test between smoking and dementia conversion did not reveal a statistically significant interaction effect. This suggests that while smoking affects DNA methylation levels in the *OXT* promoter, it does not significantly alter the relationship between methylation levels and dementia conversion in our study. Therefore, smoking’s impact on DNA methylation may be an independent (risk) factor rather than a modifier of the conversion process, reinforcing the robustness of these biomarkers regardless of smoking status.

Additionally, we identified significant effects in our analyses related to the other covariates that influenced DNA methylation levels in the *OXT* promoter including sex, age, and alcohol consumption, with notable differences between males and females. For sex, a significant increase in DNA methylation was observed in the females among all diagnostic groups when compared to the males. This strong difference in DNA methylation levels of the *OXT* promoter between males and females is not surprising; as multiple studies have demonstrated that sex can significantly affect DNA methylation patterns and epigenetic regulation in the DNA [28]. Differences in DNA methylation levels have even been observed in both healthy and diseased contexts [28, 29].

Interestingly, dementia symptomatology also varies based on sex, with females experiencing higher rates of depression, psychosis, delusions, and aberrant motor behaviors [30]. Additionally, the prevalence of developing AD dementia is twice as high in females as in males, even after accounting for their longer average lifespan [30, 31]. Furthermore, following a diagnosis of MCI, females tend to deteriorate more rapidly and exhibit faster brain atrophy compared to males [23]. Overall, these differences may be linked to sex-specific patterns in DNA methylation, which could underlie the distinct symptomatology and resilience observed between males and females. Further research is needed to specifically evaluate the role of DNA methylation changes in the *OXT* promoter within this context.

A strong association between aging and a decrease in DNA methylation levels was also observed among all diagnostic groups, an effect which appeared even more pronounced in females when compared to males. Specifically, among the SCD, MCI, and dementia groups, CpG sites 2 and 3 remained statically significant after multiple testing correction when both sexes were pooled. While these effects were not observed during the stratified analyses for males, for females within the MCI and dementia groups, a similar association for CpG sites 3 was identified after FDR-correction. The robustness of these associations underscores the critical role of aging in influencing DNA methylation patterns in females, potentially contributing to increased susceptibility or progression of dementia. A link between aging and DNA methylation changes is well established [31, 32]. Global as well as region-specific changes have been observed across time and different tissue types as well as populations [32, 33]. Hence, our findings regarding the *OXT* promoter align with the extensively documented phenomenon that DNA methylation changes are linked to longevity [22].

For females, the strong associations between age and DNA methylation levels in the *OXT* promoter, combined with the significant effects of smoking, suggest that both factors may play a role in dementia progression. Notably, a consistent decrease in DNA methylation levels within the *OXT* promoter was observed within the females when comparing trends across the different diagnostic stages, dementia converters, and in relation to aging and smoking. This indicates that both aging and smoking are linked to DNA hypomethylation in this locus, and that this reduction is seen across various stages of dementia, and within MCI females that will develop dementia. This suggests that DNA hypomethylation in the *OXT* promoter might be a common feature related to dementia progression in females, particularly in the context of aging and smoking.

In contrast, for males, the analyses highlighted nominally significant effects related to smoking and alcohol consumption, of which only the association at CpG site 4 with alcohol consumption remained FDR significant within the MCI group (*i.e.* within the dementia converters and non-converters). As mentioned earlier, age did not show significant associations with DNA methylation levels in the *OXT* promoter in males during the stratified analyses, and the primary predictors of diagnosis, and conversion to dementia also did not reveal significant effects. These results suggest that while certain covariates such as alcohol consumption may influence DNA methylation levels in the *OXT* promoter in males, the impact of the other covariates such as smoking is less pronounced and less consistent compared to females. The lack of significant associations between DNA methylation levels in this locus and factors such as diagnosis, conversion to dementia, age, and the weaker effects of smoking and alcohol consumption may suggest that methylation levels in this gene might be influenced by other factors not covered in our study.

The differential impact of our main predictors and the covariates on DNA methylation levels in the *OXT* promoter between sexes underscores the complexity of the relationship between epigenetic changes, life(style) factors, and progression of dementia. Furthermore, these findings underscore the importance of considering sex-specific and life(style) factors when studying epigenetic modifications and dementia. Our results suggest that age, smoking, alcohol consumption, and possibly other life(style) factors interact with DNA methylation levels in complex ways that may vary between sexes, potentially impacting the risk and progression of dementia. These insights highlight the need for further research to explore how these factors interplay and to refine analyses of DNA methylation levels in relation to the disease. Longitudinal studies will be necessary to fully understand their role and potential dynamic change over time.

To conclude, our results offer valuable insights into the potential prognostic utility of DNA methylation levels in the *OXT* promoter in blood, particularly for female MCI patients transitioning to dementia. The distinct findings related to dementia conversion in males and females highlight the need for further investigation into the role of DNA methylation levels in the *OXT* promoter, particularly given its potential as a prognostic tool in blood. Future research should focus on validating these findings in larger, more diverse dementia cohorts and exploring the underlying biological mechanisms driving this association.

## Ethics statement

The BioBank Alzheimer Center Limburg (BBACL) study protocol was approved by the local ethics committee (METC 15-4-100) at Maastricht University Medical Center + (MUMC+) and all participants provided written informed consent.

## Data Availability

All data produced in the present study are available upon reasonable request to the author

---

**Supplementary Table 1.**
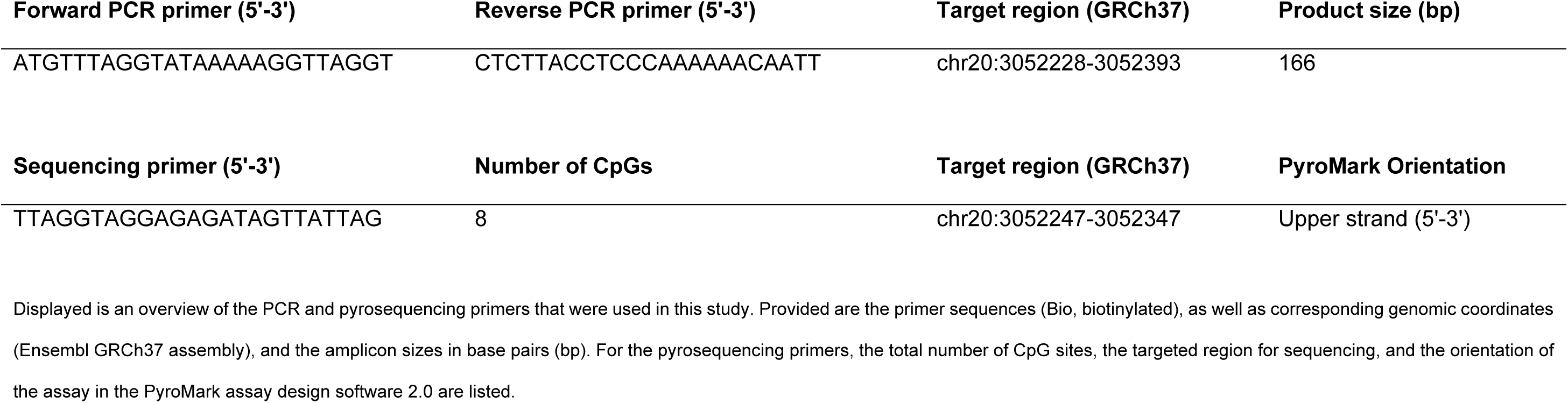
Polymerase chain reaction (PCR) and pyrosequencing primers for the *OXT* promoter.

**Supplementary Figure 1.**
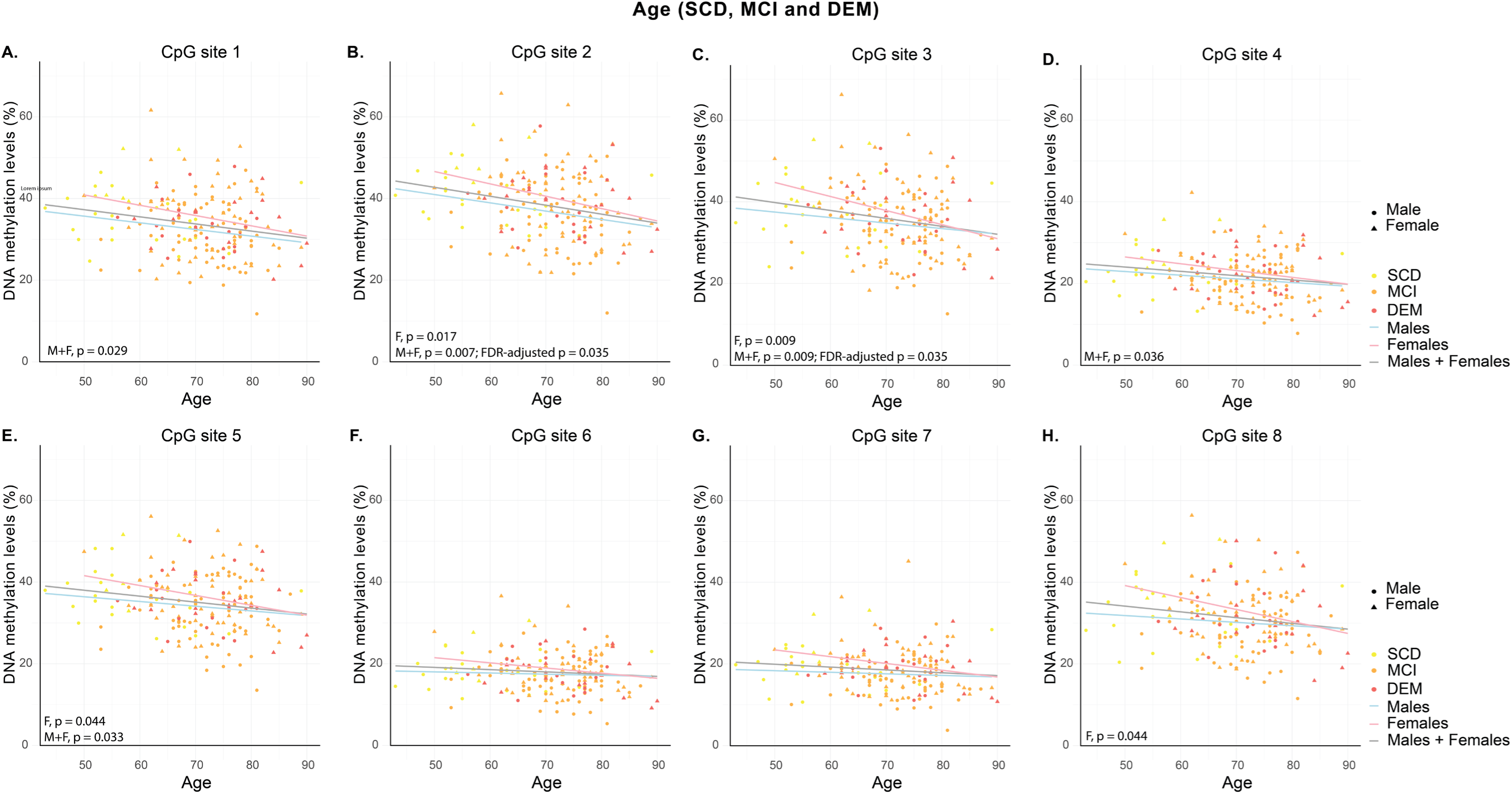
DNA methylation levels by diagnosis and sex over age for SCD, MCI, and dementia. The scatter plots demonstrate the DNA methylation levels in the *OXT* promoter per cytosine-phosphate-guanine (CpG) site for the diagnostic groups subjective cognitive decline (SCD), mild cognitive impairment (MCI), and dementia (DEM). Males and females are indicated by circles and triangles, respectively. Trend lines represent the average DNA methylation levels for the respective CpG site for males and females combined, as well as separately, over age. Nominal significance related to the covariate age is indicated with *p* < 0.05, whereas FDR-adjusted statistical significance, if applicable, is indicated with FDR-adjusted *p* < 0.05. M, male; F, female; M+F, male and female.

**Supplementary Figure 2.**
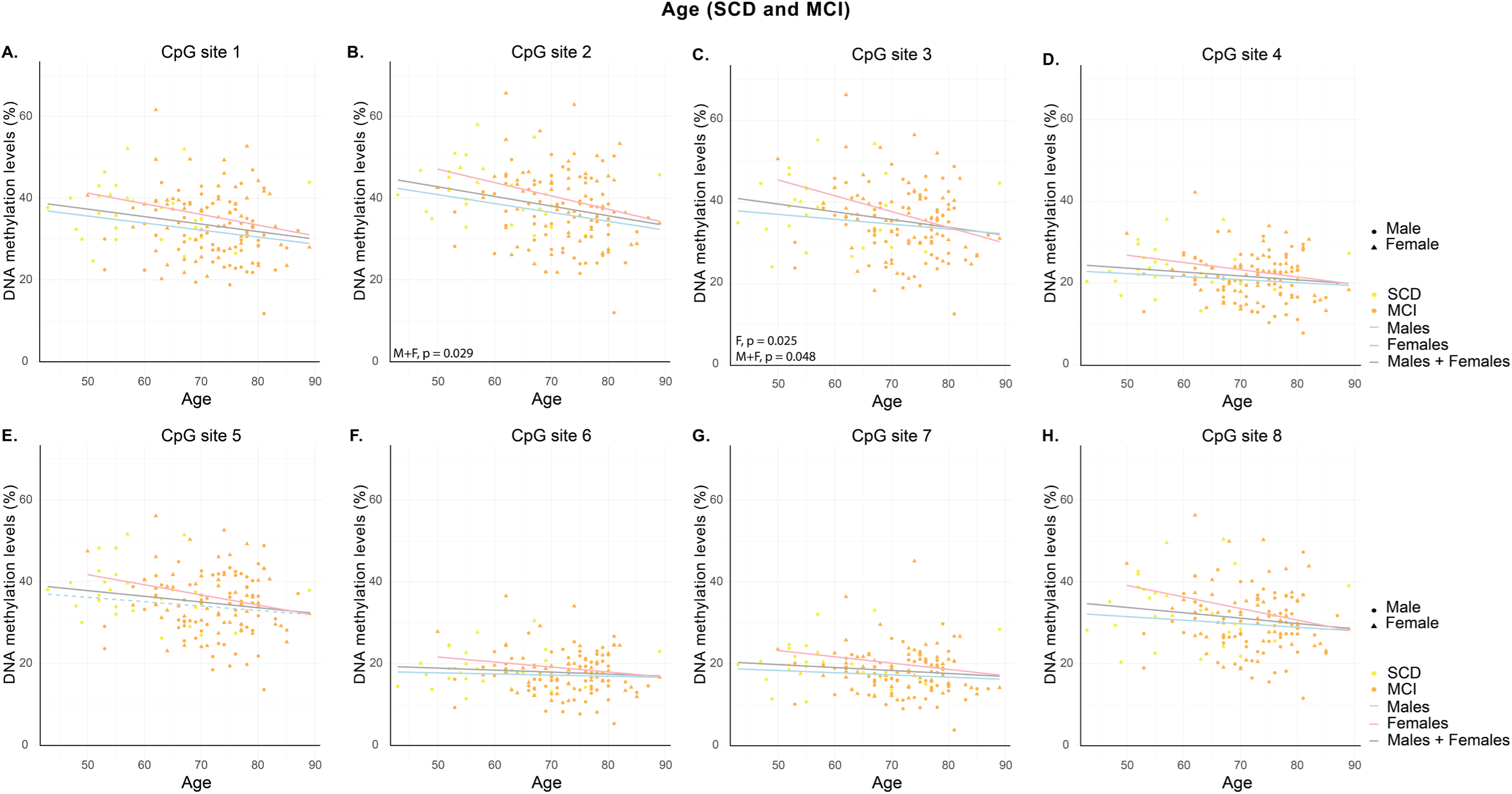
DNA methylation levels by diagnosis and sex over age for SCD and MCI. The scatter plots demonstrate the DNA methylation levels in the *OXT* promoter per cytosine-phosphate-guanine (CpG) site for the diagnostic groups subjective cognitive decline (SCD) and mild cognitive impairment (MCI). Males and females are indicated by circles and triangles, respectively. Trend lines represent the average DNA methylation levels for the respective CpG site for males and females combined, as well as separately, over age. Nominal significance related to the covariate age is indicated with *p* < 0.05, whereas FDR-adjusted statistical significance, if applicable, is indicated with FDR-adjusted *p* < 0.05. M, male; F, female; M+F, male and female.

**Supplementary Figure 3.**
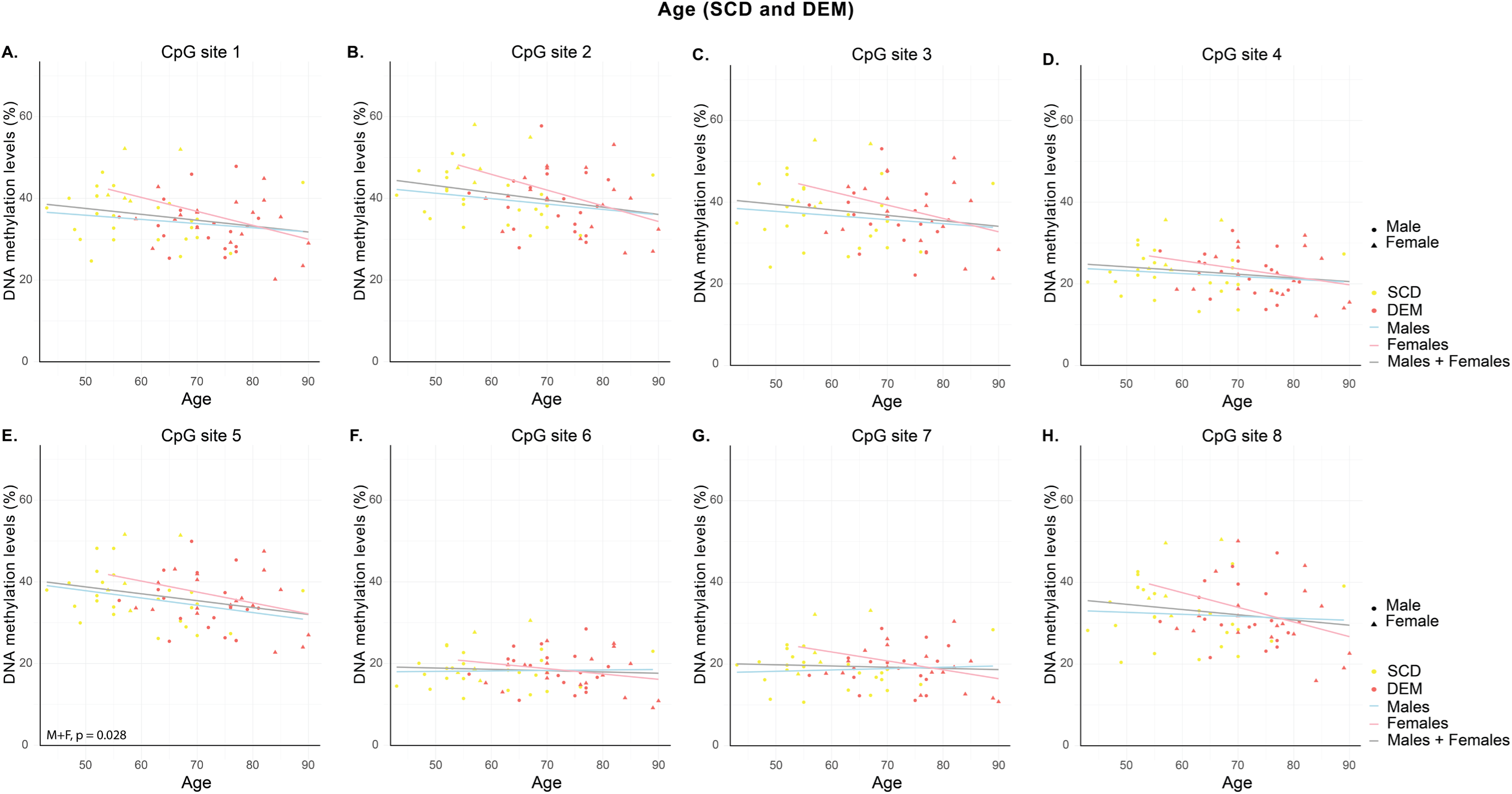
DNA methylation levels by diagnosis and sex over age for SCD and dementia. The scatter plots demonstrate the DNA methylation levels in the *OXT* promoter per cytosine-phosphate-guanine (CpG) site for the diagnostic groups subjective cognitive decline (SCD) and dementia (DEM). Males and females are indicated by circles and triangles, respectively. Trend lines represent the average DNA methylation levels for the respective CpG site for males and females combined, as well as separately, over age. Nominal significance related to the covariate age is indicated with p < 0.05, whereas FDR-adjusted statistical significance, if applicable, is indicated with FDR-adjusted p < 0.05. M, male; F, female; M+F, male and female.

**Supplementary Figure 4.**
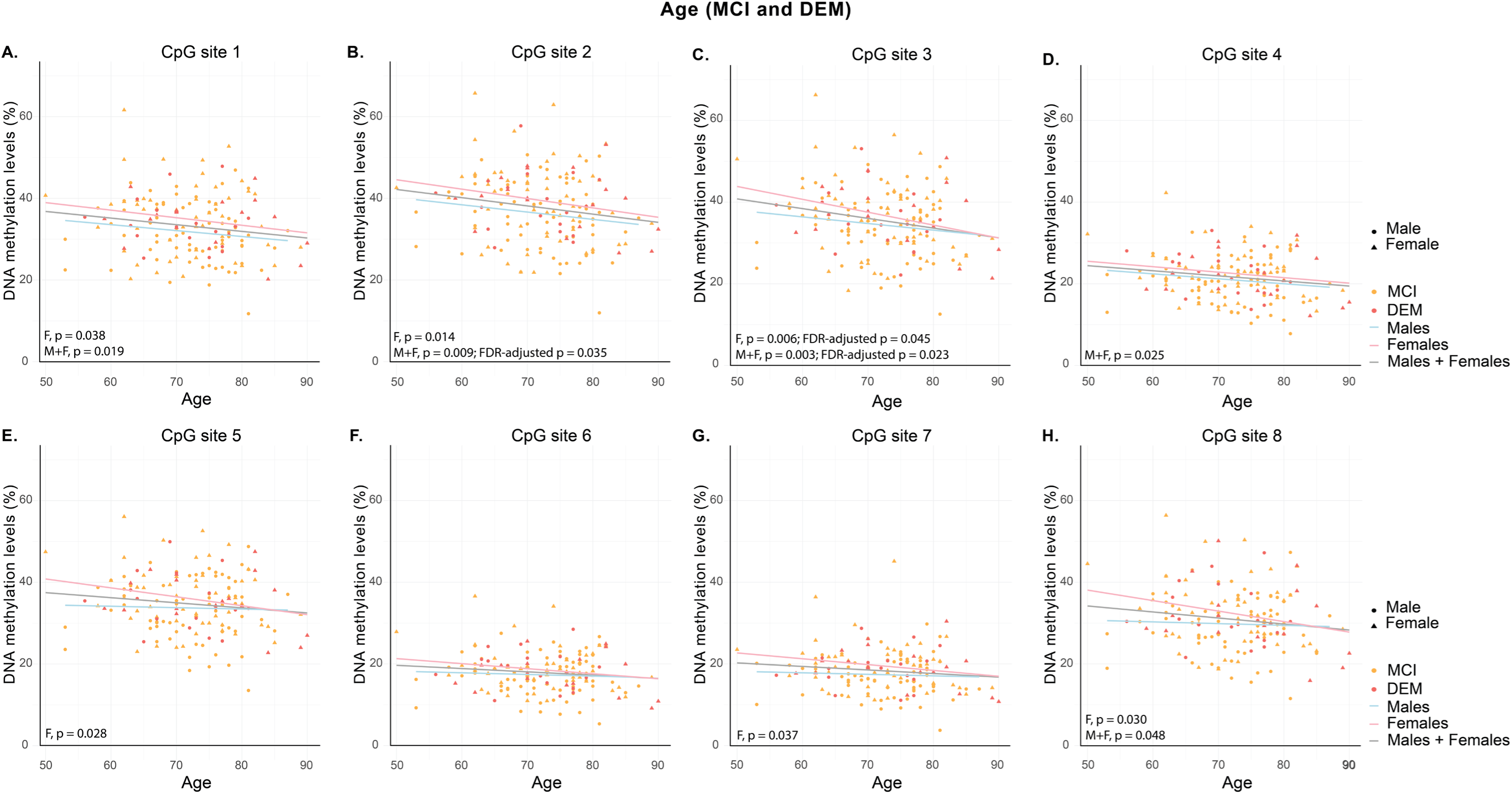
DNA methylation levels by diagnosis and sex over age for MCI and dementia. The scatter plots demonstrate the DNA methylation levels in the *OXT* promoter per cytosine-phosphate-guanine (CpG) site for the diagnostic groups mild cognitive impairment (MCI) and dementia (DEM). Males and females are indicated by circles and triangles, respectively. Trend lines represent the average DNA methylation levels for the respective CpG site for males and females combined, as well as separately, over age. Nominal significance related to the covariate age is indicated with *p* < 0.05, whereas FDR-adjusted statistical significance, if applicable, is indicated with FDR-adjusted *p* < 0.05. M, male; F, female; M+F, male and female.

**Supplementary Figure 5.**
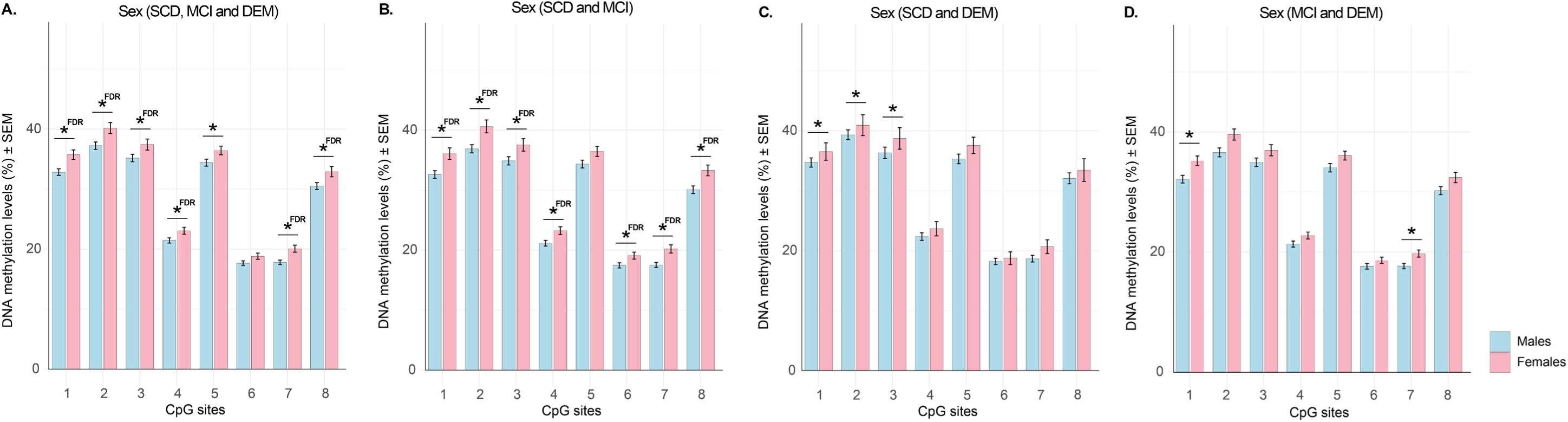
Average CpG site DNA methylation levels by sex across diagnostic contrasts. The bar charts show the average cytosine-phosphate-guanine (CpG) site DNA methylation levels in the *OXT* promoter ± the standard error of the mean (SEM). These averages are presented for males and females across the following diagnostic contrasts: subjective cognitive decline (SCD), mild cognitive impairment (MCI), and dementia (DEM); SCD and MCI; SCD and DEM; and MCI and DEM. Nominal significance (*p* < 0.05) related to the covariate sex is indicated with an asterisk (*), whereas FDR-adjusted statistical significance (FDR-adjusted *p* < 0.05), if applicable, is indicated with an asterisk followed by “FDR” (*FDR).

**Supplementary Figure 6.**
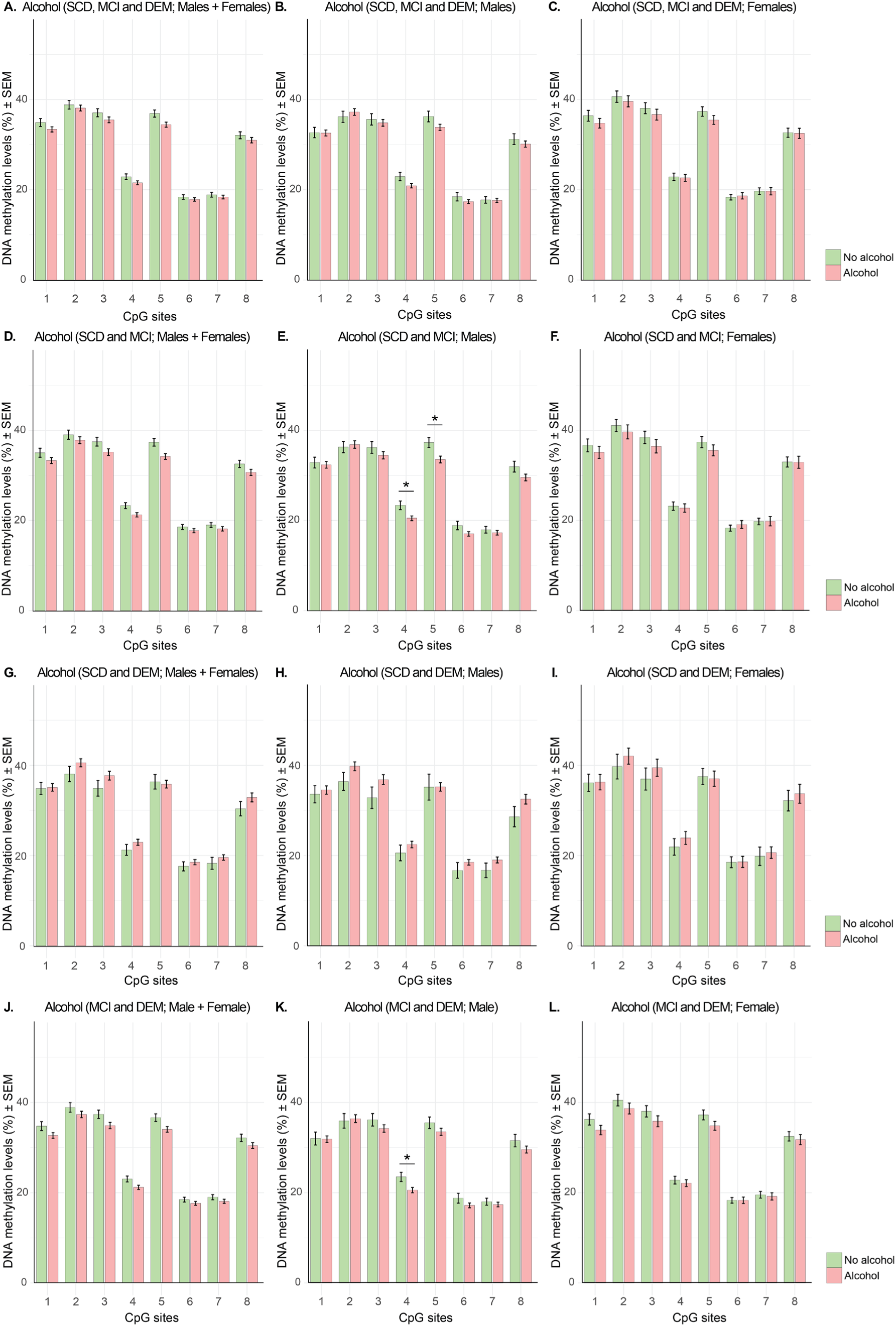
Average CpG site DNA methylation levels by alcohol consumption status across diagnostic contrasts. The bar charts show the average cytosine-phosphate-guanine (CpG) site DNA methylation levels in the *OXT* promoter ± the standard error of the mean (SEM). These averages are presented for alcohol consumers and non-consumers across the following diagnostic contrasts: subjective cognitive decline (SCD), mild cognitive impairment (MCI), and dementia (DEM); SCD and MCI; SCD and DEM; and MCI and DEM. The averages are displayed for both males and females combined, as well as separately. Nominal significance (*p* < 0.05) related to the covariate alcohol consumption is indicated with an asterisk (*), whereas FDR-adjusted statistical significance (FDR-adjusted *p* < 0.05), if applicable, is indicated with an asterisk followed by “FDR” (*FDR).

**Supplementary Figure 7.**
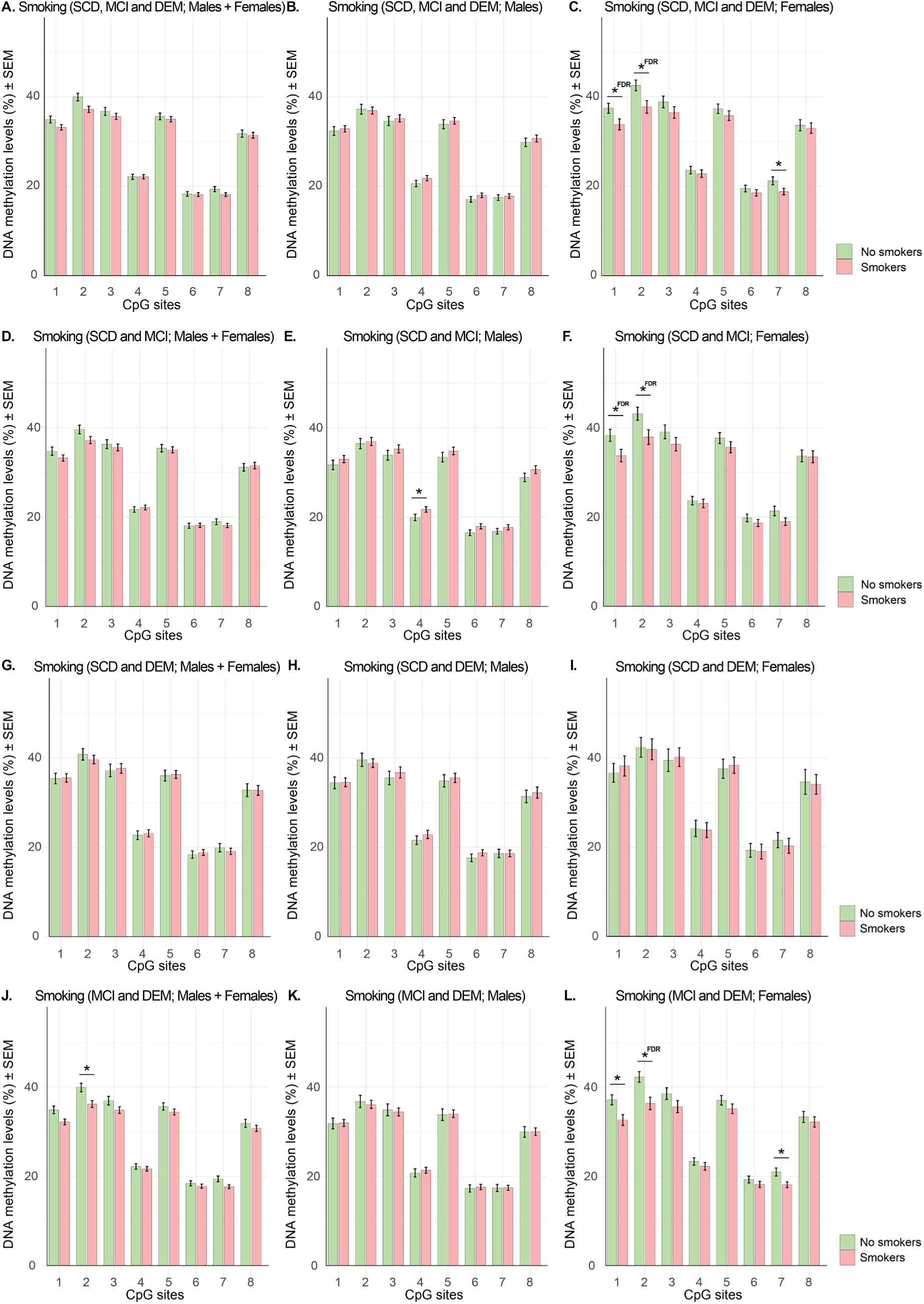
Average CpG site DNA methylation levels by smoking status across diagnostic contrasts. The bar charts show the average cytosine-phosphate-guanine (CpG) site DNA methylation levels in the *OXT* promoter ± the standard error of the mean (SEM). These averages are presented for smokers and non-smokers across the following diagnostic contrasts: subjective cognitive decline (SCD), mild cognitive impairment (MCI), and dementia (DEM); SCD and MCI; SCD and DEM; and MCI and DEM. The averages are displayed for both males and females combined, as well as separately. Nominal significance (*p* < 0.05) related to the covariate smoking is indicated with an asterisk (*), whereas FDR-adjusted statistical significance (FDR-adjusted *p* < 0.05), if applicable, is indicated with an asterisk followed by “FDR” (*FDR).

**Supplementary Figure 8.**
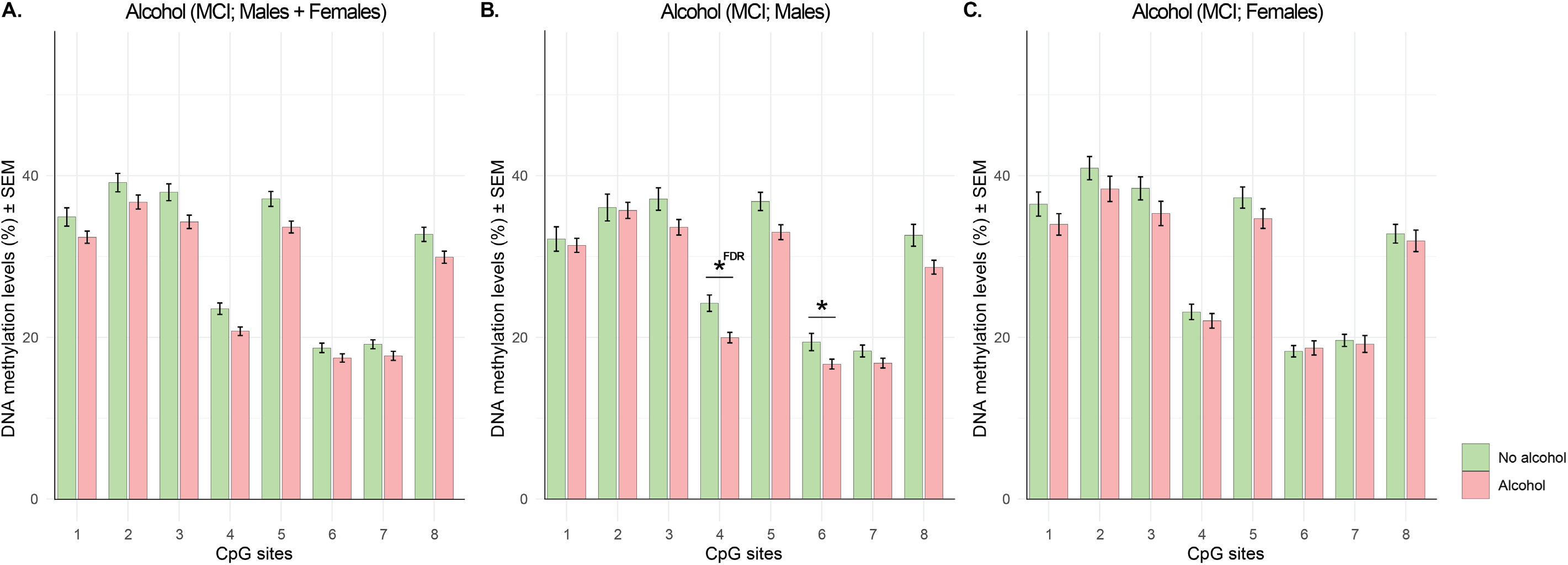
Average CpG site DNA methylation levels by alcohol consumption status across dementia converters and non-converters. The bar charts show the average cytosine-phosphate-guanine (CpG) site DNA methylation levels in the *OXT* promoter ± the standard error of the mean (SEM). These averages are presented for alcohol consumers and non-consumers across the dementia converters and non-converters in the mild cognitive impairment (MCI) group. The averages are displayed for both males and females combined, as well as separately. Nominal significance (*p* < 0.05) related to the covariate alcohol consumption is indicated with an asterisk (*), whereas FDR-adjusted statistical significance (FDR-adjusted *p* < 0.05), if applicable, is indicated with an asterisk followed by “FDR” (*FDR).

**Supplementary Figure 9.**
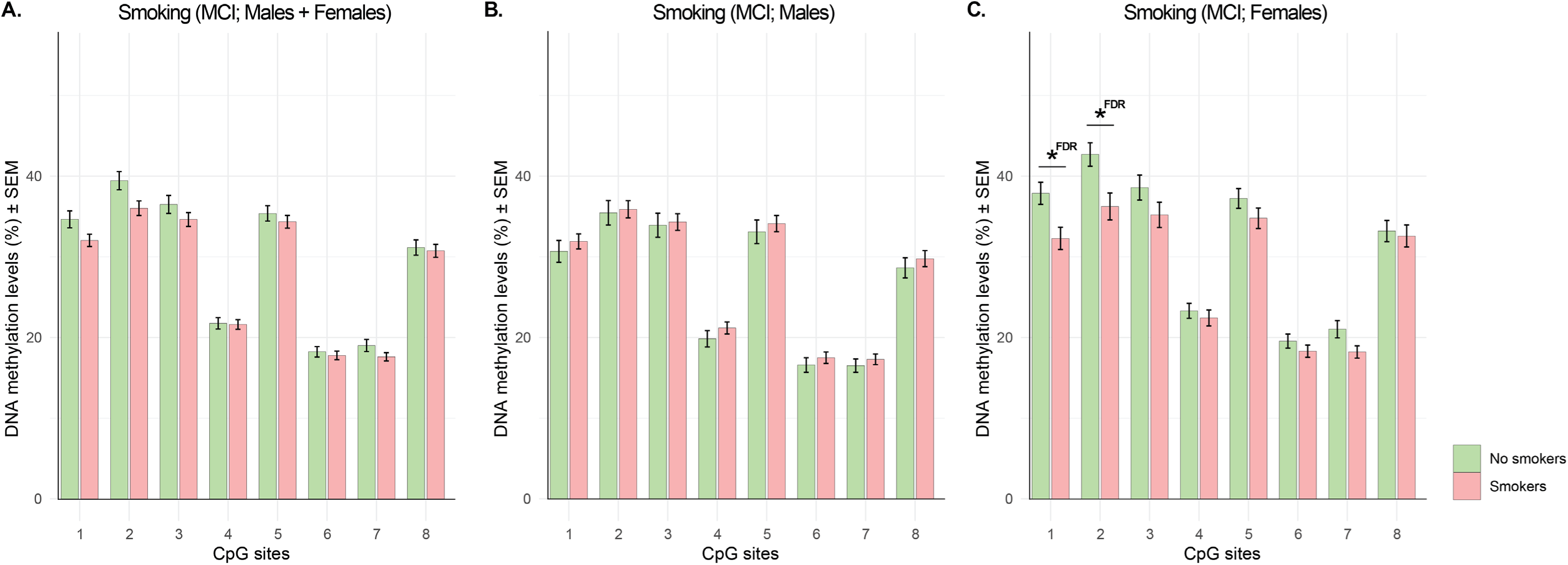
Average CpG site DNA methylation levels by smoking status across dementia converters and non-converters. The bar charts show the average cytosine-phosphate-guanine (CpG) site DNA methylation levels in the *OXT* promoter ± the standard error of the mean (SEM). These averages are presented for smokers and non-smokers across the dementia converters and non-converters in the mild cognitive impairment (MCI) group. The averages are displayed for both males and females combined, as well as separately. Nominal significance (p < 0.05) related to the covariate smoking is indicated with an asterisk (*), whereas FDR-adjusted statistical significance (FDR-adjusted p < 0.05), if applicable, is indicated with an asterisk followed by “FDR” (*FDR).

